# Nursing students satisfaction level Regarding Clinical learning environment in Peshawar

**DOI:** 10.1101/2023.01.08.23284312

**Authors:** Ihsanur Rahman, Umar Sahib

## Abstract

**Background:** of this study is to know about the students’ satisfaction level regarding clinical learning environment.

**Methodology:** A descriptive cross-sectional study design was conducted in Peshawar, KPK. Participants were selected from different semester by using random sampling technique. The data was collected through modified questionnaire.

**Results:** A total of 140 nursing students of BSN and post RN participated in which male were 54 (38.6%) and female were 86 (61.4%). The average age of the study participants was 24.41 years with a SD 5.194. The total male participants were 54, among 3 (5.5%) of them were dissatisfied from their clinical environment, 46 (85.1%) were satisfied and 5 (9.2%) were highly satisfied from their clinical environment. On the other hand among total female participant 86, among 2 (2.3%) of them were dissatisfied, 72 (83.7%) were satisfied and 12 (13.9%) participants were highly satisfied from their clinical environment.

**Conclusion:** The current study conducted in two nursing colleges, focused on satisfaction level regarding clinical environment. The result showed that the satisfaction level was in ascending order as the junior students were least satisfied. Most satisfied students were from senior most classes. The study shows the need for further research work on factors affecting satisfaction level in order to help the low level of satisfaction amongst junior classes’ students.

## Introduction

Nursing is a evidence based profession and the clinical environment is one of the most valuable and integral component in the BS nursing program (1). Clinical placement is important because it allows the nursing students to understand clinical practice, and facilitate to attain the knowledge (2). The clinical learning environment is an essential part in nursing education and has major influence on the students’ learning (3). Several definition have been presented but in this study clinical environment includes everything that surrounds the students and affect their professional development, and knowledge skills in the clinical setting (4). Additionally, the clinical learning environment can have play pivotal role in the development of the attitude, knowledge and the capability of problem solving in nursing (5). The excellence of clinical learning environment can be influenced by different factors such as ward atmosphere, leadership style of the ward manager, supervisory relationship and principles of nursing care and practice of learning in the ward (6).

It has been experienced by researchers that nursing students were not satisfied during their clinical exposure (7). Literature revealed that nursing students have clearly identified that clinical environment is not learnable as compared to class room environment, because the lecturer has more experienced as compared to clinical instructor, as the study observed that clinical instructor is fresh graduated and inexperienced, so that can influence the students learning satisfaction (8). The key aim of the study is to evaluate nursing students’ satisfaction level regarding their clinical learning environment.

## Methodology

### Study Design and Setting

A quantitative cross-sectional study design was used. The study was comported in two private nursing colleges in Peshawar. The total population was 188, while the response rate was 140. A total of 140 BS nursing students participated in this study.Sample size was calculated by using Rao soft software by taking 95 % of confidence interval and 5 % margin of error. Students were assessed through adopted questionnaire.

### Ethical consideration

Permission letter was signed from the Director Admin of the two nursing colleges for data collection. Informed consent was taken from each participant. Confidentiality and anonymity of the participants were maintained.

### Tools

The 3 likert scale (dissatisfied, satisfied and highly satisfied) adopted questionnaire was used on the following different domains, ward atmosphere, learning environment in the ward and student teacher relationship.

## Results

A total of 140 nursing students of BSN and Post RN participated in study; male were 54 (38.6%) and female were 86 (61.4%). The mean age of the students was 24.41 (SD 5.194) years. The students from BSN were 109 (77.9%) and post RN was 31 (22.1%). The student’s satisfaction level was categorized into three levels, highly satisfied whose score is greater than 90, satisfied with score of 40 - 90, dissatisfied whose score less than 40 and the total score was 105. According to their clinical rotation, the last units they had attended. Students had the clinical placement of participants in 29 (20.7%) were from ICU, 51 (36.4%) from general ward, 9 (6.4%) were from pediatric and 49 (35.0%) participants were from others and 2 participants have not mentioned their clinical placement as shown in table 1.

**Table 1.** Demographic characteristics

|  |  |
| --- | --- |
| Total participants | 140 |
| **Gender** |  |
| Male | 54 |
| Female | 86 |
| **Year of Study** |  |
| Year 1 (combined BSN+PRN) | 19 |
| Year 2 (combined BSN+PRN) | 40 |
| Year 3 (BSN) | 35 |
| Year 4 (BSN) | 46 |
| **Clinical Placement** |  |
| ICU | 29 |
| general ward | 51 |
| pediatric ward | 9 |
| Other | 49 |
| **Satisfaction Level** |  |
| Satisfied | 118 |
| highly satisfied | 17 |
| Dissatisfied | 5 |

Among all, 5 (3.6%) participants were dissatisfied, while satisfied were 118 (84.3%), more over 17 (12.1%) students were found to be highly satisfied from their clinical learning environment. The satisfactions level of the participants from their clinical learning environment as shown in the figure1.

**Figure 1.**
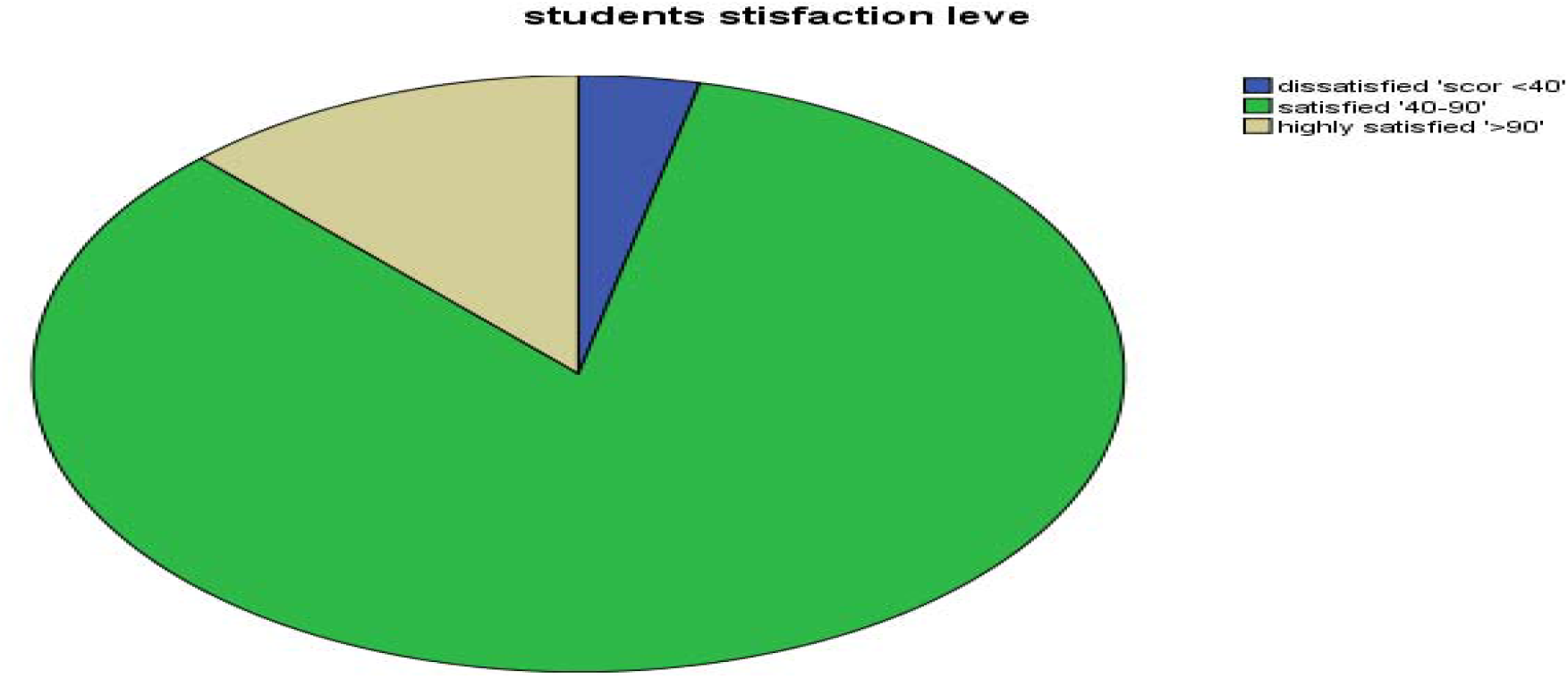
Satisfaction level of the clinical environment

Chi-square test applied to identify association between student’s satisfaction level with the variables such as gender, year of study, clinical placement and program. However, gender and program were not found to be significant.

Association between students satisfaction level and the participants year of the study was significant (P=0.017). Students of year 4 BSN were more satisfied (93.47%), year 3 were less satisfied (88.57%), year 2 were lesser (77.5%) satisfied while students of year 1 post RN (68.42 %) were least satisfied as shown in table 2.

**Table 2.**
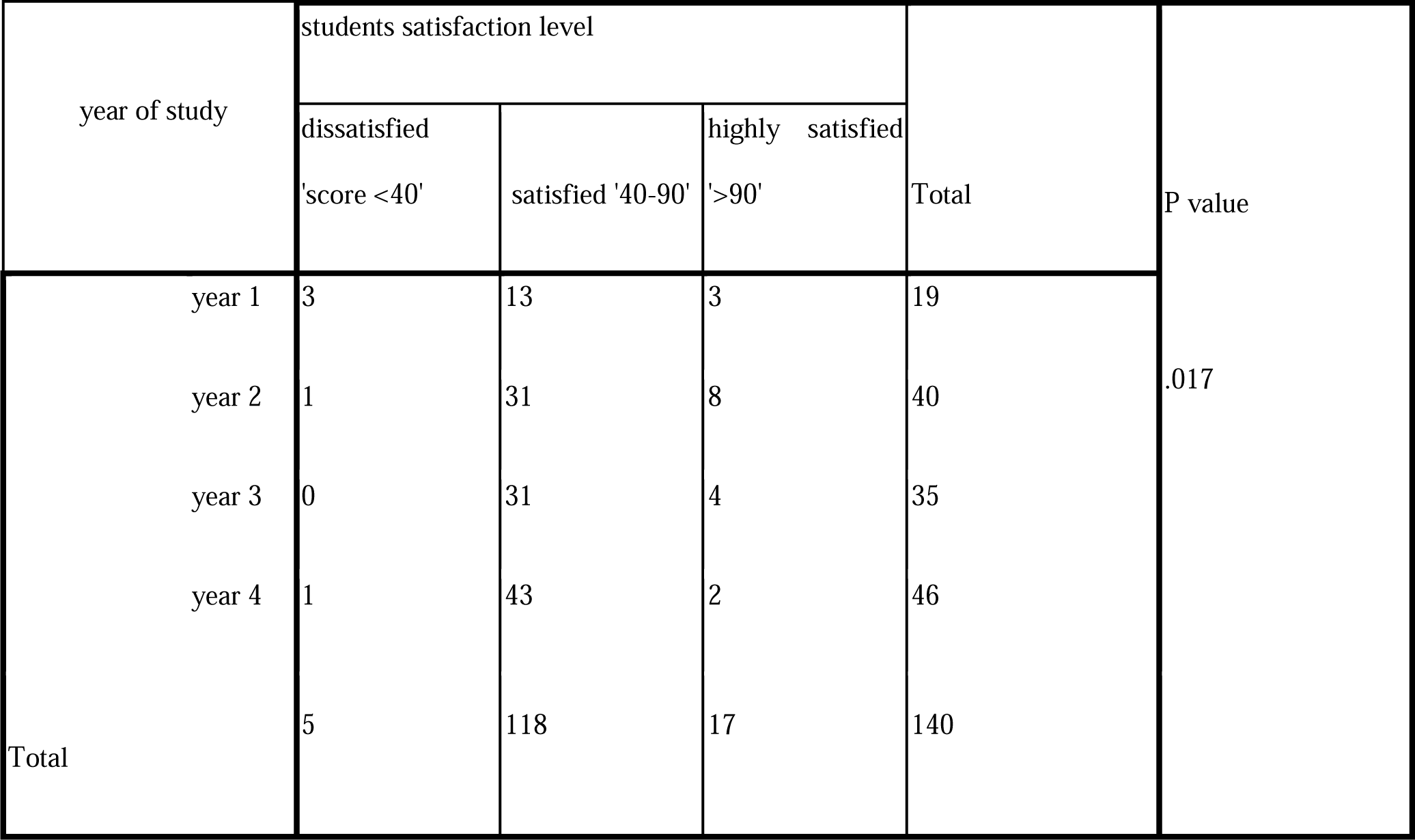
satisfaction level according to the years

Association between clinical placement and students satisfaction level was also found to be significant (P=0.034). last clinical placement in general ward participants were more satisfied (88.23%), others were least satisfied (85.71%), ICU participants were less satisfied (68.96%) and pediatrics ward all were satisfied (100%) as shown in table 3.

**Table 3.** satisfaction level according to the clinical placement

| Clinical placement | students satisfaction level | Total |
| --- | --- | --- |

|  | dissatisfied<br>'score <40' | satisfied '40-90' | highly satisfied<br>'>90' |  | P value |
| --- | --- | --- | --- | --- | --- |
| ICU | 4 | 20 | 5 | 29 | .034 |
| General ward | 0 | 45 | 6 | 51 |  |
| Pediatrics ward | 0 | 9 | 0 | 9 |  |
| Other | 1 | 42 | 6 | 49 |  |
| Total | 5 | 116 | 17 | 138 |  |

## Discussion

This study has conducted in two private nursing colleges at Peshawar, KP, Pakistan; large numbers of the students were satisfied with their clinical learning environment. The findings are supported by other studies (2,9). The reason for good satisfaction level of students from their clinical learning environment is good facilitation of students in private institutions, because such institutions do not compromise on their standards as they have to compete with other colleges in the market. Study could have brought different results if we would have included other college’s especially public sector colleges.

Another reason for nursing students’ satisfaction from clinical learning environment could be good profile of teaching faculty. Generally, private colleges offer handsome salaries for which they hire most competent faculties. Consequently, faculties of private institutions keep better learning environment for their students. The current study shows that majority of the satisfied students were from the senior class, it might be, they are more exposed to clinical environment, more confident, had more command on clinical skills and possessed rich knowledge as compare to junior students. On contrary, a study conducted in Cyprus (10) reported that the junior most students were more satisfied from their clinical learning environment. The difference in findings might be due to availability of mentors to the junior students in Cyprus. Therefore, it is highly recommendable to apply the practice of mentorship in other countries as well. It is also worth mentioning that in the current study, the least satisfied (68.42%) participants were from the Post RN BSN first year. A study has found that Post RN BScN students had comparatively less critical thinking and less level of knowledge; therefore their comprehension about clinical environment could be comparatively weaker than generic students (11). Also, another reason for their least satisfaction from clinical environment could be that all the post RN (BScN) students who participated in this study were doing jobs and classes in same days and they could sphere lesser time to get themselves adjusted in the clinical environment.

## Conclusion

The study is about the nursing student’s satisfaction level regarding clinical learning environment. As a whole (12.1%) were highly satisfied, (84.3%) were satisfied, and (3.6%) were dissatisfied. In nutshell, majority of the students were satisfied from their clinical learning environment. The satisfaction level was in ascending order as the junior students were least satisfied. Most satisfied students were from senior most classes. The study shows the need for further research work on factors affecting satisfaction level in order to help the low level of satisfaction amongst junior classes’ students.

Strength of this study was first study conducted in Peshawar to explore nursing students’ satisfaction from their clinical learning environment. Probability sampling technique was applied, which increases the generalizability of findings on private colleges. Weakness of the study we include private colleges in our study. We could not include the public colleges for generalizability. As in this study the junior students were found to be least satisfied, therefore it is recommended that the clinical management must assign mentor to the junior students in order to help them adjust to the clinical environment.

## Supporting information

ERB

## Data Availability

data will be available upon the request of the publisher

## Acknowledgment

None

