## Supplementary material for "Nursing students satisfaction level Regarding Clinical learning environment in Peshawar": ERB

### KHYBER MEDICAL UNIVERSITY

**KMU-IPHSS Ethics Board**


No. KMU/IPHSS/Ethics/2022/AB/073 Dated: 24-10-2022

### TO WHOM IT MAY CONCERN

Certified that Ethical Approval has been granted to the project title

**Nursing student's satisfaction level regarding clinical environment in Peshawar**

submitted by Ihsanur Rehman, Scholar of Master in Public Health (MPH), under the supervision of Dr. Naveed Sadiq, Assistant Professor, IPH&SS, Khyber Medical University, Peshawar.

**Director (IPH&SS)**
IPH&SS-Ethics Board
